# Cytogenetic signatures of recurrent pregnancy losses

**DOI:** 10.1101/2020.07.01.20144535

**Authors:** Svetlana. A. Yatsenko, Cristina Quesada-Candela, Devereux N. Saller, Stacy Beck, Ronald Jaffe, Stefan Kostadinov, Judith Yanowitz, Aleksandar Rajkovic

## Abstract

**Objectives:** To investigate the incidence of chromosomal abnormalities in the products of conception (POC) of patients with spontaneous miscarriages (SM) and with recurrent pregnancy losses (RPL), and to determine biological mechanisms contributing to RPL.

**Design:** Retrospective cohort study.

**Setting:** University-affiliated medical center.

**Patients:** During a 20-years period, 12,096 POC samples underwent classical chromosome analysis as a part of standard clinical care.

**Interventions:** Cytogenetic findings were classified into six categories and compared between the SM and RPL cohorts.

**Main Outcome Measures:** RPL-specific cytogenetic abnormalities and sex bias in POCs with autosomal aneuploidy.

**Results:** Analysis of a large cohort of RPL patients has identified an increased incidence of inherited and *de novo* structural chromosome abnormalities, recurrent polyploid conceptions, and complex mosaic alterations. These abnormalities are the signature of genomic instability, posing a high risk of genetic abnormalities to offspring independent of maternal age. Predominance of male conceptions in the RPL cohort points toward X-linked etiology and gender-specific intolerance for certain genetic abnormalities.

**Conclusions:** Our study showed several possible genetic etiologies of RPL, including parental structural chromosome rearrangements, predisposition to meiotic nondisjunction and genomic instability in patients with karyotypically abnormal POCs. Loss of karyotypically normal fetuses might be attributed to defects in genes essential for fetal development and survival, as well as aberrations affecting the X chromosome structure or function. Molecular studies of parental and POC genomes will help to identify inherited defects in genes involved in meiotic divisions and DNA repair to confirm our hypotheses, and to discover novel fetal-essential genes.

## INTRODUCTION

Miscarriage (SM), defined as spontaneous pregnancy loss before 20 weeks of gestation, is a common clinical problem in the general reproductive population and is estimated to occur in 10-15% of clinically recognized pregnancies (US Department of Health and Human Services, 1982). The American Society for Reproductive Medicine (ASRM) and the World Health Organization (WHO) have defined the recurrent pregnancy losses (RPL) as the loss of two or more pregnancies (documented by ultrasonography or histopathologic examination), not necessarily consecutive [1,2]. The prevalence of RPL varies among reports based on differences in the definitions and criteria used, however all the studies recognize RPL as an important reproductive health issue, affecting approximately 2-5% of couples of a reproductive age based on ASRM criteria, with a current increasing trend [1,3,4]. The potential etiologies for RPL described are: genetic factors (random chromosomal abnormalities during gametogenesis, unbalanced chromosomal aberrations resulting from parental balanced rearrangements, postzygotic embryonic aneuploidy, gene mutations affecting fetal viability); endocrine factors (thyroid dysfunction, diabetes mellitus, polycystic ovary syndrome); autoimmune factors such as Antiphospholipid Antibody Syndrome (APS); fetal and placental infections; chronic exposure to teratogenic agents; anatomical factors (congenital uterine malformations, acquired anatomic disorders); cervical incompetence; and environmental, occupational, and personal habits [5].

Cytogenetic or chromosomal abnormalities are the most common cause of miscarriage during the first trimester. It is estimated that 50% of miscarriages are due to chromosomal abnormalities, which mainly arise *de novo* in the embryo from parents with normal karyotypes [6,7]. The high incidence of age-related meiotic errors during oogenesis makes advanced maternal age an important risk factor for early miscarriage [8]. The majority of sporadic losses result from random numerical or structural chromosome rearrangements or triploidy, with a low recurrence risk in women younger than 35 years of age. Approximately 2% of first trimester miscarriages are caused by tetraploidy, which is thought to result from a failure to undergo cytokinesis during the first or a subsequent mitotic cell division in the zygote [9]. Maternal age appears to have little effect on the occurrence of such defect, however relatively little is known about the genetic causes of fetal tetraploidy.

In contrast to spontaneous miscarriages, a much smaller proportion of women with recurrent pregnancy losses have karyotypically abnormal conceptions [10–13]. Recurrent causes include chromosomal abnormalities derived from a parental balanced rearrangement. Unbalanced structural alterations inherited from phenotypically normal carrier parents are independent of maternal age, with a constant unchanging risk for each conception [14]. It is estimated that 2% to 5% of couples with RPL have a structural chromosomal abnormality, as opposed to 0.2% of the general population [15, 16]. The type of rearrangement and whether the male or female partner is the carrier is crucial in determining the reproductive risk and the outcome of the pregnancy. In general, the frequency of aneuploidy in RPL patients is similar to a group with spontaneous miscarriages. In RPL patients, maternal age does not always predict risk of an aneuploid pregnancy [17]. In a subset of RPL women, rates of aneuploid conceptuses are significantly higher than the average for their given age. Such losses are commonly labeled as random events, however molecular mechanisms governing genetic predisposition to aneuploidy remain to be elucidated. The majority of recurrent miscarriages are associated with normal karyotype, and do not have a clearly defined etiology.

In the present study we evaluated and compared cytogenetic abnormalities found in the products of conception (POC) from spontaneous miscarriages and RPL with the goal to deduce biological mechanisms contributing to fetal pathology and recurrent fetal losses and to understand molecular etiologies that may explain the losses of karyotypically normal fetuses.

## MATERIALS AND METHODS

### Study population

In this retrospective study, we examined the findings of karyotype analysis from POC specimens referred to the Pittsburgh Cytogenetics Laboratory during a 20-year period. The study was approved by the University of Pittsburgh Institutional Review Board (20060192). Patients were categorized into two cohorts: recurrent pregnancy losses (RPL), including patients with two or more consecutive miscarriages; and a cohort with non-recurrent fetal loss, comprising the patients with a single miscarriage (SM). Chromosome analysis was attempted on a total of 12,096 POC samples, of which no culture growth was observed in 1,366 samples (11.3%) and 10,730 were successful. Among successful karyotypes, 5,728 (47.4%) yielded normal results, 2,891 with a normal 46,XX karyotype and 2,837 with a normal 46,XY karyotype (Table 1). The rest, 5,002 (41.4%) of POCs had an abnormal karyotype. Among our POC specimens, the ratio of 46,XY to 46,XX results is 1:1.02 (2837:2891), which is statistically not different from the expected 1:1, indicating a low possibility of maternal cell contamination.

**Table 1.**
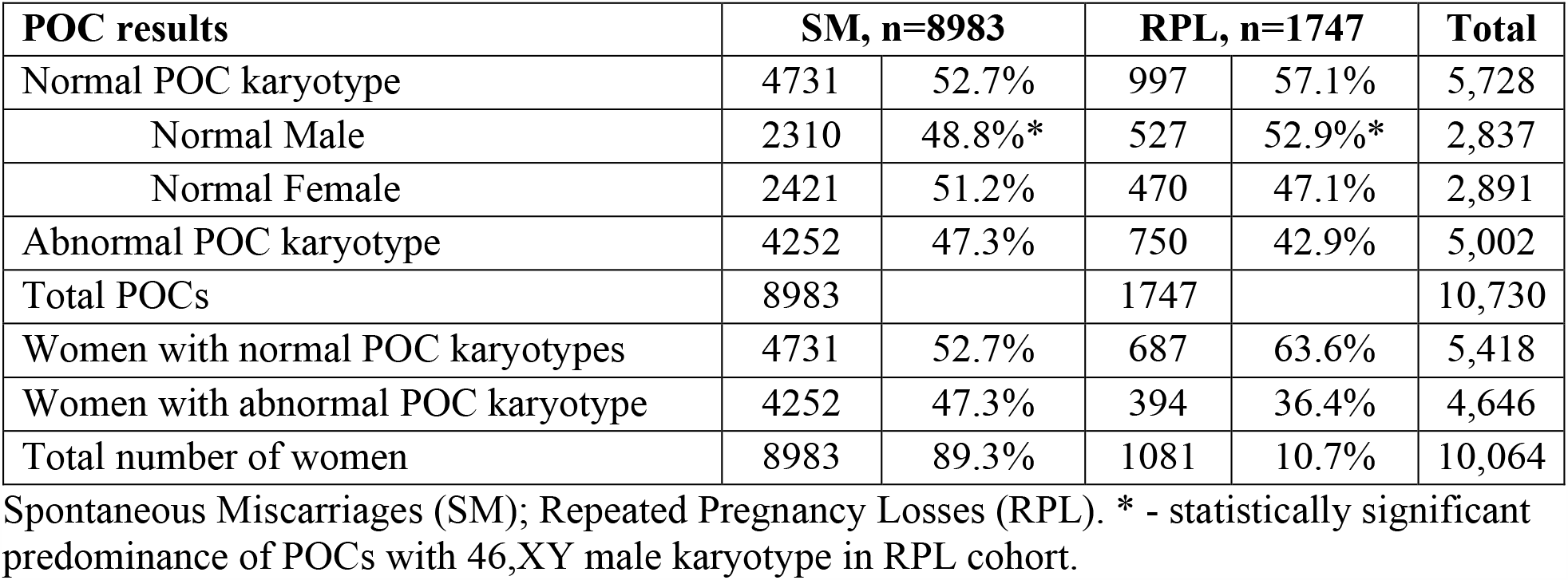
Summary of cytogenetic findings in the study cohorts.

| **POC results** | **SM, n=8983** |  | **RPL, n=1747** |  | **Total** |
| --- | --- | --- | --- | --- | --- |
| Normal POC karyotype | 4731 | 52.7% | 997 | 57.1% | 5,728 |
| Normal Male | 2310 | 48.8%* | 527 | 52.9%* | 2,837 |
| Normal Female | 2421 | 51.2% | 470 | 47.1% | 2,891 |
| Abnormal POC karyotype | 4252 | 47.3% | 750 | 42.9% | 5,002 |
| Total POCs | 8983 |  | 1747 |  | 10,730 |
| Women with normal POC karyotypes | 4731 | 52.7% | 687 | 63.6% | 5,418 |
| Women with abnormal POC karyotype | 4252 | 47.3% | 394 | 36.4% | 4,646 |
| Total number of women | 8983 | 89.3% | 1081 | 10.7% | 10,064 |

Spontaneous Miscarriages (SM); Repeated Pregnancy Losses (RPL). \* - statistically significant predominance of POCs with 46,XY male karyotype in RPL cohort.

A total 8,983 POCs were from women who experienced a single spontaneous miscarriage, 4731 (52.7%) of them had normal karyotype. Cytogenetic studies were performed on 1,747 POC samples from 1,081 women with RPL (Table 1). Among RPL cases, 57.1% (997/1747) POCs from 63.6% (687/1081) women had normal karyotype. Among women with normal POC findings, the mean age in the SM cohort was 30.2 years, and 31.9 years in RPL patients, ranging in age from 18.4 to 45 years at the time of miscarriage.

Our study has focused on SM and RPL cohorts with abnormal karyotypes: 4,252 POCs in SM cohort, and 750 POCs from 394 women in RPL group (Table 1). The RPL study cohort included 63 women with at least three pregnancy losses, and 331 patients with at least two documented pregnancy losses. Abnormal POC karyotypes were observed from a total of 4,646 women. Among women with abnormal POC findings, the mean age of patients was 33.5 years in the cohort of SM patients, and 35.1 years in the cohort of RPL patients. Out of 394 RPL patients with abnormal findings, 29.4% (318/1081) had two or more POC samples with abnormal karyotype. Women with an abnormal karyotype in a single POC sample and normal findings for other losses were accounted for 7% (76/1081) of total RPL cases.

### Chromosome analysis by the G-banded karyotype

Tissue obtained from fetal organs, chorionic membranes, umbilical cord and placental villi were cultured, harvested and analyzed by conventional G-banding cytogenetic analysis. Specimens that contained only maternal decidua were rejected from chromosome analysis and are not included in this study. Overall, ∼62% of analyses were performed on fetal tissues along with chorionic membranes or placental villi, and in ∼38% of samples placental villi were analyzed as fetal tissue was not available or did not grow.

### Statistical Analysis

To determine if a certain type of abnormality is more or less likely to be carried by a person in the RPL group than in the SM group, we performed one- and two-tailed Chi-square tests at the 95% confidence level for the equality of proportions between two cohorts. In order to analyze sex differences for particular aneuploidies in the products of conception, Pearson Chi-square test was applied. The objective was to test how likely the observed distribution of data fitted with distribution that is expected. In the case of males and females the expected distribution would be 50% each. Categorical variables were presented as n(%) and values p< 0.05 were accepted as statistically significant.

## RESULTS

### Chromosomal abnormalities in POC samples

We classified karyotypic alterations into six different categories (Table 2) based on a) the presence of structural chromosomal aberration in a diploid karyotype, b) aneuploidy involving an autosome, c) aneuploidy involving sex chromosomes, d) triploid conceptions, e) POCs with a tetraploid karyotype, and f) complex karyotypes comprising multiple abnormalities. We compared a relative frequency of aberrations between the SM and RPL groups (Figure 1A). We also evaluated the frequencies of structural chromosome rearrangements, aneuploidy for each autosome, and mosaicism for multiple cell lines among each category in patients with RPL versus SM cohorts. Out of 394 RPL patients, 318 had abnormal cytogenetic results on two or more miscarried fetuses. Among women with abnormal POC cytogenetic findings, 25/318 (7.8%) had parental chromosome aberrations, 24/318 (7.5%) had recurrent polyploid conceptions, and 14/318 (4.4%) patients had 2 abortuses each with aneuploidy for the same chromosome (trisomy 16 (n=5), monosomy X (n=3), trisomy 22 (n=2), and trisomies 6, 7, 10, 21 (n=1 each). We observed a single case of chimerism with a karyotype discordant for sex chromosomes 46,XY/47,XX,+15.

**Table 2.** Summary of karyotypic abnormalities in patients with a single spontaneous Miscarriages (SM) and Repeated Pregnancy Losses (RPL).

| **Karyotype category** | **SM, n=4252** |  | **RPL, n=750** |  |
| --- | --- | --- | --- | --- |
| Diploid | 132 | 3.1% | 44 | 5.9% |
| Aneuploid | 2436 | 57.3% | 451 | 60.1% |
| Sex chromosome abnormalities | 539 | 12.7% | 58 | 7.7% |
| Triploid | 751 | 17.7% | 117 | 15.6% |
| Tetraploid | 184 | 4.3% | 36 | 4.8% |
| Complex | 210 | 4.9% | 44 | 5.9% |
| Number of patients | 4252 | 91.5% | 394 | 8.5% |

**Figure 1.**
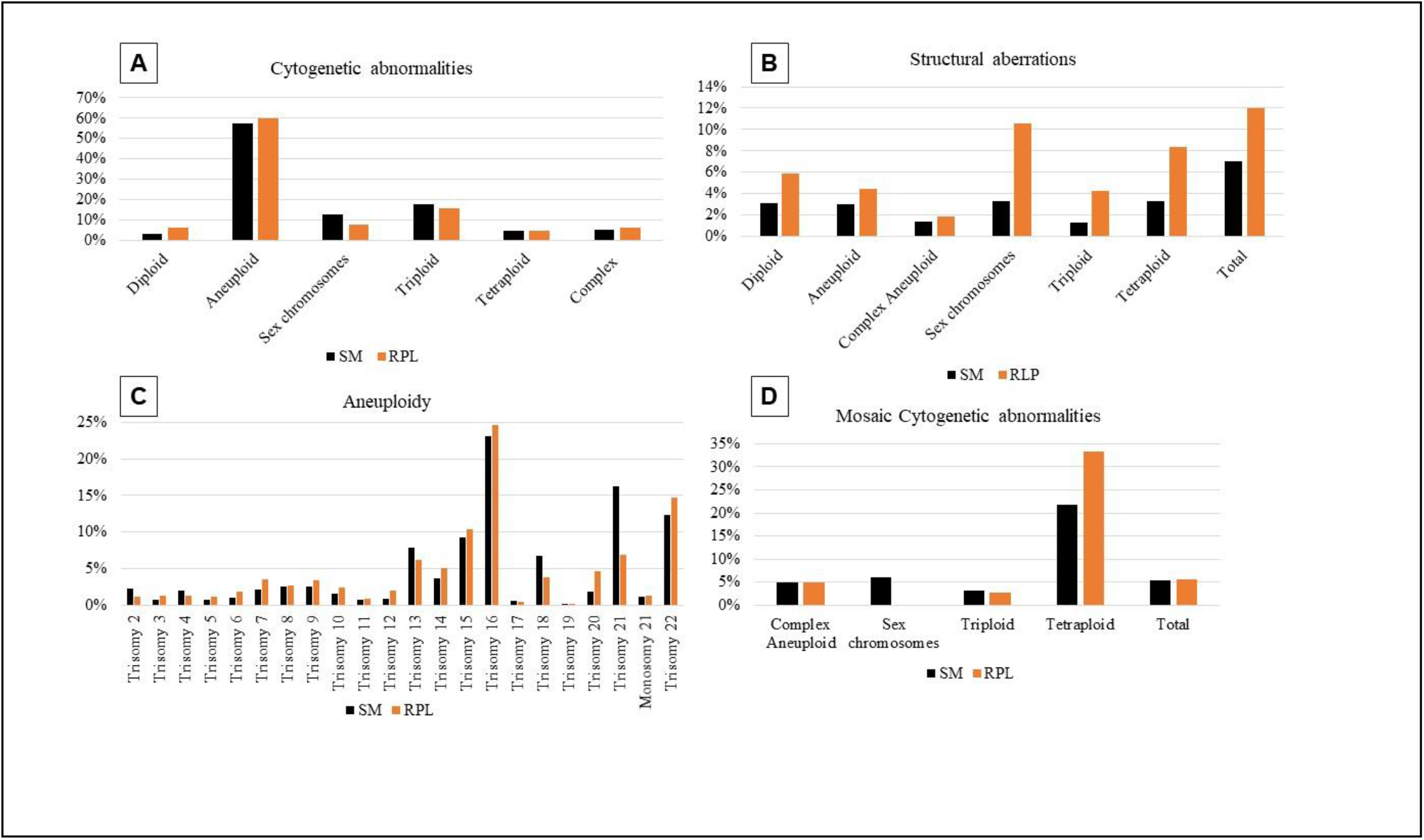
The rate of cytogenetic alterations in POC samples from cohorts of patients with a spontaneous miscarriage (SM) and women with recurrent pregnancy losses (RPL). **(A)** Histogram showing percentage of samples with abnormalities in each category: diploid karyotypes with structural chromosomal aberration; autosomal aneuploidy; sex chromosome aneuploidy; triploidy; tetraploidy; and complex abnormalities. **(B)** Percentage of structural chromosome abnormality observed among cytogenetic categories. **(C)** Rate of aneuploidy for each chromosome. **(D)** Frequency of mosaic cytogenetic alterations.

### Prevalence of structural chromosome abnormalities in RPL

Structural rearrangements were observed in 6.6% (230/4252) of POCs from SM group, including 1.2% (50/4252) unbalanced reciprocal translocations (0.6% inherited), and 1.2% (52/4252) unbalanced karyotypes due to Robertsonian translocations. Structural abnormalities were observed in 12.1% (91/750) of POC samples (6.3% inherited) from the RPL cohort (Figure 1B), with the prevalence of unbalanced reciprocal translocations rearrangements seen in ∼5.1% of POC samples, which is at least 4-folds more frequent (p<1.57E-05) in comparison to the SM cohort (Table 3). In the RPL cohort, recurrent structural rearrangements were observed in POCs from 6.5% (25/394) of patients, or in 53/750 POC samples, 11% of which had karyotypes due to 3:1 segregation and 17% of POC samples also had aneuploidy for a chromosome not involved in a rearrangement. Parental testing was available on ∼94% of couples with POC carrying a reciprocal unbalanced translocation, showing the mother as a carrier in ∼48% and the father in ∼32% of cases, while both parents had normal karyotypes in 19% of cases. There was no difference between SM and RPL cohorts for incidence of other structural rearrangements, including marker, ring, and isodicentric chromosomes, deletions, inversions, duplications, although detection of some aberrations might be limited by the resolution of G-banded chromosome analysis. The rate of Robertsonian translocation was slightly higher in RPL cohort, accounting for 2.1% of abnormal karyotypes, versus 1.6% seen in SM group. Outcomes of Robertsonian translocation were not included in the above calculations, but instead were placed in the aneuploidy category. Parents from the RPL cohort have an overall significantly higher rate of structural chromosome rearrangements among all karyotypic groups, including cases when structural rearrangements were present along with triploid and tetraploid karyotypes (Figure 1B).

**Table 3.** Structural chromosome abnormalities in SM and RPL cohorts.

| **Structural abnormality** | **SM, n=4252** |  | **RPL, n=750** |  |
| --- | --- | --- | --- | --- |
| Unbalanced reciprocal translocations | 50 | 1.2% | 38 | 5.1% |
| Robertsonian translocations | 52 | 1.2% | 16 | 2.1% |
| Balanced translocations | 43 | 1.0% | 13 | 1.7% |
| Isodicentric chromosomes | 28 | 0.7% | 7 | 0.9% |
| Marker chromosomes | 24 | 0.6% | 6 | 0.8% |
| Deletions | 57 | 1.3% | 6 | 0.8% |
| Inversions | 19 | 0.4% | 3 | 0.4% |
| Duplications | 4 | 0.1% | 2 | 0.3% |
| Ring chromosomes | 5 | 0.1% | 0 | 0.0% |
| Total | 282 | 6.6% | 91 | 12.1% |

### Aneuploidy and chromosomal mosaicism among RPL

We investigated the incidence of autosomal aneuploidies in POC samples to determine the differences between SM and RPL cohorts (Figure 1C). Trisomies 16, 21, 22, 15, 13, and 18 were the most common detected with frequencies of 23.1%, 16.2%, 12.3%, 9.3%, 7.9%, and 6.9%, respectively. Overall, aneuploidy is equally common in both groups. Interestingly, trisomy for chromosomes 21 and 18 has been less frequently observed in RPL patients, in 6.9% (p<0.00001) and 3.8% (p< 0.001), respectively. The same observation was true for sex chromosome aneuploidy observed in 7.7% (p< 3.552E-07) in RPL versus 12.7% in SM (Figure 1A). These findings indicate that trisomy 21, 18, and monosomy X are not the major contributors to a recurrent miscarriage. Chromosomal mosaicism, the presence of two or more populations of genetically different cells, has been documented in ∼5.5% of POC samples (Figure 1D). The most common mosaic aneuploidies were trisomy 16, 13, 2, and 22, identified in 0.35%, 0.35%, 0.28%, and 0.21% of aneuploid POC, respectively. The major differences between the SM and RPL cohorts were observed in the frequency of mosaic findings in the samples with a tetraploid karyotype (Figure 1D). In the RPL cohort, we detected a much higher rate (p<0.035) of mosaicism for an additional tetraploid cell line(s) with a gain or loss of one or multiple chromosomes, aneuploid cell line and a tetraploid cell line resulted from a doubling of genome seen in aneuploid cells, additional structural aberrations in a tetraploid cells, tetraploid cells along with normal diploid cells. These findings indicate a failure in postzygotic cell division processes and genomic instability among RPL patients.

No differences between SM and RPL cohorts were observed for the cases with double aneuploidy. Extra chromosome 21 was the most common abnormality present in ∼41% of cases with double aneuploidy, followed by trisomy 16 (23%) and trisomy 14 (13%) in a combination with trisomy for other chromosomes.

### Age dependence of chromosomal findings

Chromosome aberrations are detrimental to fetal development and survival, and commonly are the cause of spontaneous abortions. We evaluated the frequency of karyotypic abnormalities by age with the goal to determine if RPL patients have an age-independent increase for certain alterations (Figure 2). The mean age in our RPL cohort with abnormal karyotypic findings is 35.1 years, with two peaks at ages 34 and 40 years, which most likely reflects the time of RPL diagnosis with multiple prior fetal losses (Figure 2A). The most common abnormalities we detected in POC samples of RPL patients at age 34 were the unbalanced structural chromosome rearrangements, explained by parental balanced alterations (Figure 2B) and sex chromosome structural abnormalities (Figure 2E). These findings are also consistent with the observation of an age-independent increase for structurally abnormal chromosomes in POCs from the RPL cohort (Figure 2I).

**Figure 2.**
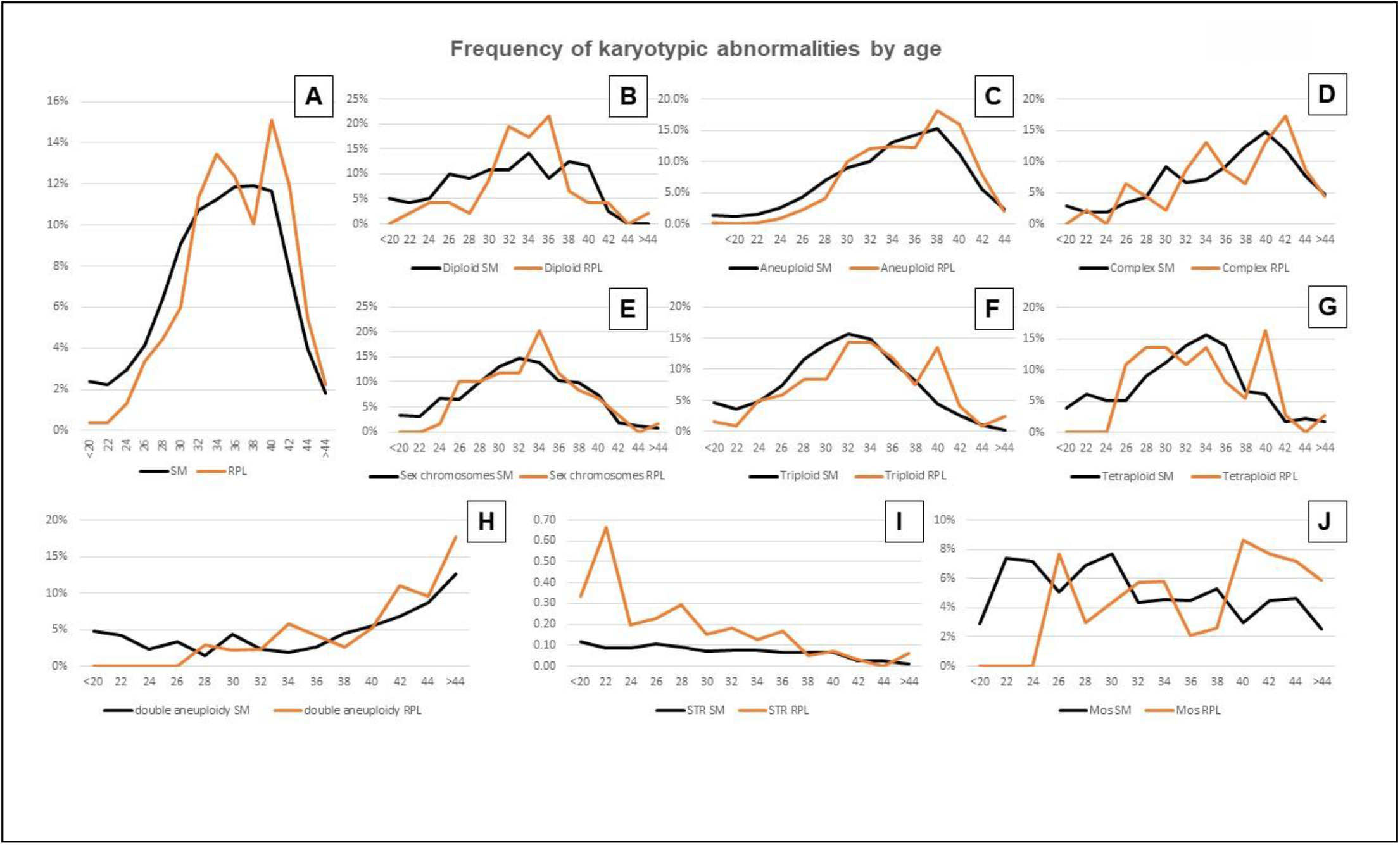
Frequency of karyotypic abnormalities by maternal age at the time of miscarriage. **(A)** Age distribution in patients with abnormal POC findings. **(B)** Percentage of diploid karyotype with structural chromosome abnormality in patients of different age. **(C)** Rate of aneuploidy by age. **(D)** Frequency of complex karyotype with multiple alterations in age groups. **(E)** Rate of sex chromosome aneuploidy by age. **(F)** Frequency of triploidy. **(G)** Rate of tetraploid karyotypes in patients of different age. **(E)** Rate of double aneuploidy (aneuploidy for two or more chromosomes) with age. **(E)** Overall frequency of structural chromosome rearrangement in patients of different age. **(E)** Rate of mosaic findings in POC samples by mothers’ age.

Interestingly, a number of RPL patients in our cohort had recurrent losses affected by various structural rearrangements: deletions, duplications, translocations, and formation of abnormal chromosomes, suggesting a predisposition to chromosomal breakage and defects in DNA repair. We also observed a higher rate of 3:1 chromosome missegregation events involving parentally-inherited structurally abnormal chromosomes, categorized as complex alterations (Figure 2D). At age 40 years, RPL patients had a higher rate of aneuploid and triploid pregnancies (p<0.0048), as well as complex chromosomal abnormalities (Figure 2C, F). The frequencies for trisomy 12 (2.0%, p< 0.05) and trisomy 20 (4.7%, p< 0.0047) were significantly higher in RPL patients, as compared to 0.9% for trisomy 12 and 1.8% for trisomy 20 in the SM cohort. The complex alterations after age 40 are likely due to a cumulative effect of age-related risk for aneuploidy and genetic predisposition to chromosome rearrangement, which has been also observed for samples with aneuploidy for two or more chromosomes (double aneuploidy, Figure 2H). Mosaic findings are also more common after age 40 in RPL patients (Figure 2J). Remarkably, the rate of tetraploid POCs in the RPL cohort is increased in patients of all ages (Figure 2G).

### Impact of autosomal aneuploidy onto male and female fetuses

We also investigated potential fetal sex differences in trisomy pregnancies. From a total number of 2885 aneuploid products of conception the frequency of males (M) and females (F) were 51.3% and 48.7%, respectively. The observed male to female ratio in not statistically different from the expected 1:1 The most common aneuploidies in both males and female POCs from RPL patients were trisomies 16 (M= 21.5%; F= 23.8%), 21 (M= 14.9%; F= 13.7%) and 22 (M= 13.0%; F= 11.8%) (Figure 3). Trisomies for all the autosomal chromosomes were found with the only exception of chromosome 1. Trisomy 1 appears to affect early stages of embryonic development, resulting in implantation failure or an early fetal lethality, and therefore, is not likely to survive long enough to be seen in spontaneous abortions.

**Figure 3.**
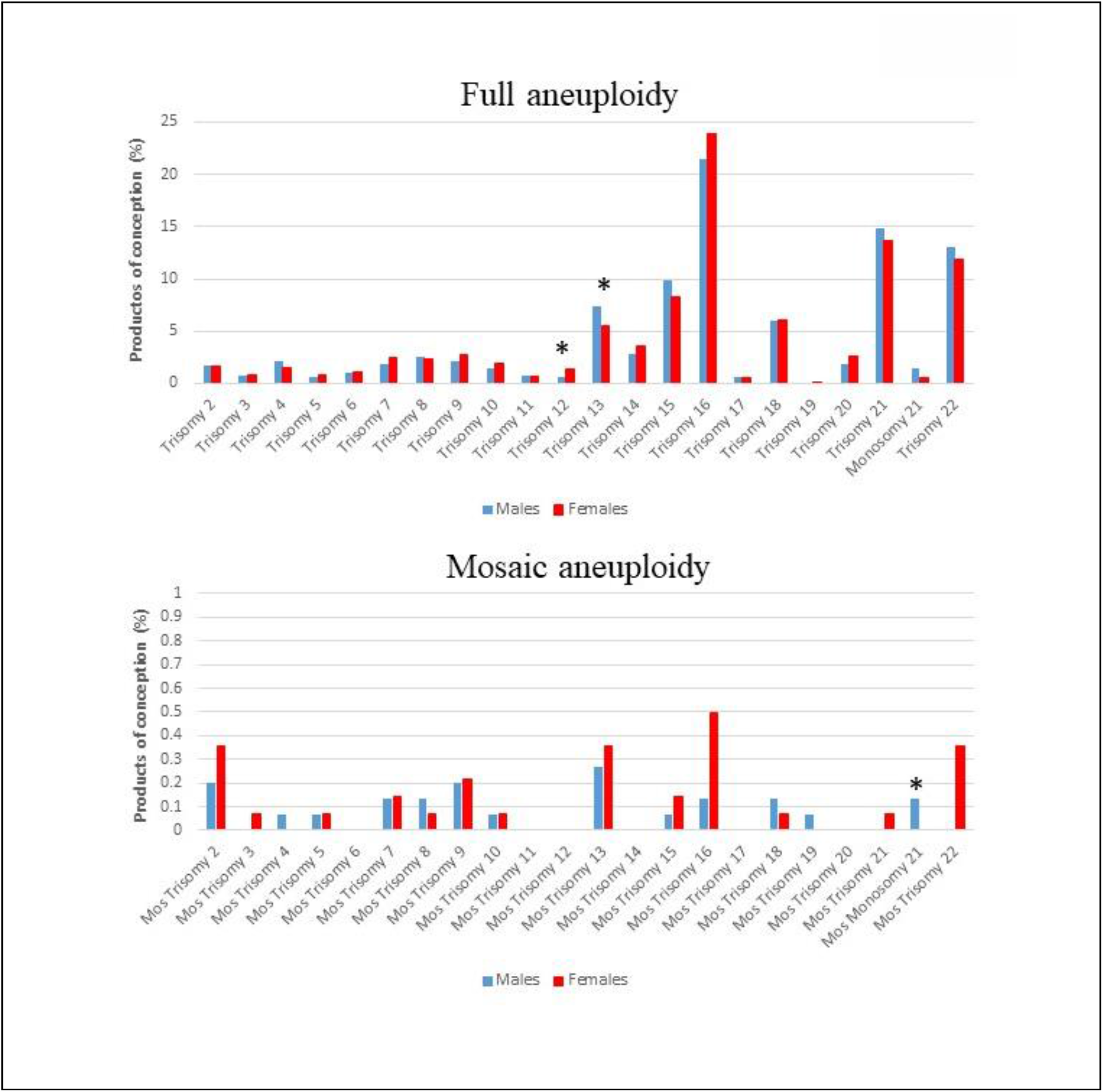
Prevalence of aneuploidy among male and female POCs. **(A)** Frequency of full chromosome aneuploidy observed in female (red bars) and male (blue bars) POCs from both SM and RPL groups. * - statistically significant sex bias. **(B)** Frequency of mosaic aneuploidy in female and male conceptions.

The majority of trisomic conceptuses, even those whose karyotype may be viable in the neonate, result in miscarriage. That is the case for full trisomy involving chromosomes 13, 18 and 21. Sex differences were found in products of conception with full trisomy 13 and trisomy 12. Full trisomy 13 was significantly more frequent (p< 0.05) in males (7.4%) than in females (5.5%), (Figure 3A). By contrast, trisomy 12 was significantly more frequent in females than in males (1.4% vs 0.6%; p < 0.05) (Figure 3A). No differences were found between males and females for trisomies 18 (M= 6.0%; F= 6.1%) and 21 (M= 14.9%; F= 13.7%) (Figure 3). Other aneuploidies appeared in the study showing a potential gender susceptibility. Despite the overall low frequency, prevalence of monosomy 21 was significantly higher in males than in females (1.5% vs 0.6, p< 0.05) (Figure 3A). These findings indicate that certain chromosomal abnormalities are less tolerated in fetuses of one gender versus another.

## DISCUSSION

In this study, we investigated abnormalities found by karyotype analysis in POC samples obtained from women with spontaneous miscarriages and a cohort of patients with recurrent pregnancy losses. Our study revealed that, in contrast to the SM group, POC from women with RPL have a much higher rate of structural chromosome abnormalities, recurrent polyploid conceptions, and mosaicism in tetraploid POCs. Remarkably, maternal age is not a risk factor for these abnormalities. The most common cause of a fetal loss during the first trimester is chromosome imbalance, which may result from a whole chromosome aneuploidy or a segmental aneuploidy due to transmission of a derivative chromosome from the parent who carries a balanced rearrangement. Reciprocal translocations were found in ∼ 6.5% of couples in our RPL cohort, consistent with the 2-8% rate reported by other studies [9,10], however the etiology of recurrent miscarriage in the majority of affected couples remains unexplained.

The risk of aneuploidy in an embryo is greatly influenced by maternal age and is well recognized as a cause of a sporadic miscarriage [18]. Aneuploidy can also be a causative factor in some couples with RPL, however multiple studies and our findings indicate a lower rate of aneuploidy in RPL cohorts compared to patients with spontaneous miscarriages [10–12]. Patients with RPL can be categorized into three subgroups: 1) with recurrent fetal losses and normal karyotypes, 2) patients with abnormal POC karyotypes, and 3) those with abnormal and normal POC findings. In our study, ∼64% of women with RPL had recurrent normal POC karyotypes. The etiology of miscarriage in this group is not likely to be associated with chromosomal alterations, and little is known about monogenic causes of RPL in chromosomally normal conceptions. Importantly, the resolution of G-banded chromosome analysis is not sufficient to reveal cryptic chromosome alterations, microdeletions, and duplications. Other limitations of karyotype analysis also include inability to detect regions with homozygosity and uniparental disomy. Genome sequencing studies have shown that in up to 12% of couples with RPL, one of the parents is a carrier of a balanced chromosomal aberration, but chromosomal rearrangements are cryptic in up to 40% of cases, and, therefore, undetectable by karyotype analysis [19]. High resolution copy number analysis may aid in detection of cryptic chromosomal imbalances in RPL [20–22].

The vast majority of RPL patients with abnormal karyotypes in multiple miscarriages and normal parental karyotypes do not exhibit the same type or the same chromosome abnormality. In our study, ∼30% of RPL patients had 2 or more chromosomally abnormal losses, POC from 4.4% of RPL women were affected by recurrent trisomy, and 7.5% patient had recurrent polyploidy in their POC samples. Although re-occurrence of chromosomal abnormality and even the re-occurrence of the same trisomy may represent a random event, it may also indicate a genetic predisposition to aneuploidy in a subset of RPL patients, particularly in young women whose aneuploidy rate is higher than expected for their age group. Recurrent trisomy for the same chromosome can be due to a parental gonadal mosaicism [23]. Gonadal mosaicism for aneuploidy can also result in depletion of ovarian reserves, which is supported by multiple studies linking unexplained RPL to a high incidence of maternal diminished ovarian reserve [24,25]. Other possible explanations include defects in genes that control spindle checkpoint, chromosome segregation, and separation of sister chromatids during meiosis. Defects in these processes can lead to a meiotic aneuploidy in oocytes, aneuploidy for multiple different chromosomes, and chromosomal mosaicism.

The high rate (7.5%) of recurrent polyploidy (triploidy and tetraploidy) in a cohort of our RPL patients is another unexpected finding. Triploidy is one of the most common chromosome abnormalities in humans, observed in ∼15-20% of all spontaneous abortions [26]. By random chance, the risk of recurrent polyploid conceptions in POC samples is estimated to be as high as 4%. The frequency of polyploid conception could be underestimated if miscarriage occurs before it can be successfully evaluated by karyotype analysis. Although rare, familial and recurrent cases of triploid conceptions strongly suggest a genetic predisposition. An oocyte’s ability to complete meiotic divisions and retain a haploid chromosome complement might be impaired by genetic alterations, such as pathogenic variants in *MEI1, REC114*, or *TOP6BL* genes [27,28], or due to other causes leading to recurrent digynic triploidy. Alternatively, oocytes may have a deficiency in the polyspermy blockage mechanisms, leading to androgenetic polyploid conceptions.

In the case of tetraploidy, we also noticed a high rate of RPL patients with a mosaicism for a diploid/tetraploid mixture and tetraploid cells containing additional aneuploidy. Cytokinetic failure at 8–16 cell stage embryos can result in the formation of multinucleated (polyploid) blastomeres in the combination with normal diploid cells. Blastomere fusion or failure of cytokinesis can be observed in >50% of aneuploid embryos [29]. Interestingly, early embryo development utilizes many biological pathways implicated in cell proliferation during tumorigenesis [30,31]. Defects in cytokinesis and tetraploidization of genome in tumor cells are also associated with genomic instability characterized by mosaicism, gains and losses of individual chromosomes. Remarkably, we observed a similar karyotypic signature characterized by mosaicism and aneuploidy for various chromosomes among POC samples in RPL cohort.

Errors during fertilization and impaired cleavage morphokinetics can lead to an embryo with an unstable genome and multiple abnormal blastomeres with chaotic chromosome abnormalities. Different risk and inherited factors may lead to embryonic chromosomal abnormalities, including patients’ genetic predisposition to meiotic and mitotic errors, defective response to DNA damage, fertilization errors and aberrant cleavage dynamics, increasing a likelihood of genomic instability in developing embryos and subsequently leading to recurrent fetal losses. The presence of chromosomal abnormality and mosaicism in POC sample may indicate a deficiency in the pathways responsible for the maintenance of genomic integrity and overall a sign of genomic instability. The underlying genetic causes responsible for these processes are yet unknown, and are the subjects of future studies.

In the RPL cohort with normal karyotypes, we observed a significantly higher ratio (p< 0.01) of the XY-containing POCs (52.9% vs 48.8% of males in the SM group with normal karyotype), which may indicate an X-linked etiology of RPL in some women. At least 6% of fertile women carry pathogenic copy number variants or point mutations in the X-linked genes associated with congenital disorders [32]. Several X-linked genes are known to cause prenatal male lethality and recurrent fetal losses, and many more remain to be discovered. The majority of X chromosome aberrations in males are submicroscopic, undetectable by karyotype analysis. Generally, the manifestations of X-linked conditions are more severe in male than female conceptions, although defects in X chromosome inactivation may result in a female-biased lethality. Sex-biased predisposition has been described among individuals affected by neurodevelopmental disorders, autoimmune diseases, some cardio-vascular defects, such as aortic aneurism, and other genetic conditions. We investigated if sex bias exists in POCs with aneuploidy for a single chromosome. Our study showed a significantly skewed male:female ratio for conceptions with trisomy 12 and trisomy 13, indicating a selection against males. Studies evaluating the survival of live born children with trisomy 13 showed that females have the lower mortality rate than males [33]. Remarkably, a few live born infants, predominantly girls, have been reported with mosaic trisomy 12 [34], consistent with our observations that male fetuses with a full or mosaic trisomy 12 do not likely to survive beyond the first trimester. The proportions of chromosome-specific trisomies are very similar between preimplantation embryos [18] and POC samples in our study, with a few exclusions. There is a much lower incidence of trisomies for chromosomes 1, 11, 17, and 19 in POCs, suggestive that conceptions of both genders with trisomy for these chromosomes are eliminated during the early stages of embryonic development.

RPL is a heterogeneous condition that may require a comprehensive genetic testing to establish an underling etiology for a given couple. Early diagnosis of RPL to enable early treatment is essential, as this condition might be complicated by age related aneuploidies and diminished ovarian reserve. Abnormal chromosomal structure and pathogenic sequence changes in genes involved in early embryonic and fetal development, as well as genes critical for the maintenance of genome integrity should be the subjects of further studies.

## CONCLUSIONS

Our study suggested a few possible molecular etiologies of recurrent miscarriage, including parental structural chromosome abnormalities, predisposition to genomic instability, meiotic nondisjunction, and fertilization errors leading to aneuploid and polyploid conceptions. We hypothesize that inherited defects in genes involved in meiotic divisions and DNA repair are the major cause of fetal loss in young patients with a high rate of chromosomal abnormalities. Loss of karyotypically normal fetuses might be attributed to a maternally inherited X-linked abnormality. Molecular studies of parental and POC genomes will help to confirm our hypothesis and discover novel genes essential for fetal development and survival.

## Data Availability

All data are available.

## ACKNOWLEDGEMENTS

We are thankful to the staff of the Pittsburgh Cytogenetics Laboratory for the technical assistance in cytogenetics studies.

